# Dynamics of the SARS-CoV-2 epidemic in the earliest-affected areas in Italy: Mass screening for SARS-CoV-2 serological positivity (SARS-2-SCREEN)

**DOI:** 10.1101/2020.06.06.20124081

**Authors:** Gabriele Pagani, Dario Bernacchia, Federico Conti, Andrea Giacomelli, Rossana Rondanin, Vittore Scolari, Patrizia Boracchi, Cecilia Eugenia Gandolfi, Silvana Castaldi, Elia Biganzoli, Massimo Galli

**Affiliations:** Department of Clinical and Biomedical Sciences (DIBIC) “L. Sacco”, University of Milan, Italy; Post graduate School in Public Health Dept Biomedical Sciences for Health, University of Milan, Italy; Fondazione IRCCS Ca’ Granda Ospedale Maggiore policlinico di Milano, Italy; Department of Clinical Sciences and Community Health & DSRC, University of Milan, Italy; Institut Curie-PSL Research University, CNRS, Sorbonne Université, UMR3664, F-75005, Paris, France; Medispa s.r.l.

**Author notes:** **Corresponding Authors:** Gabriele Pagani, ASST FBF-Sacco, Infectious Diseases Dep., III Division, Luigi Sacco Hospital, Via G.B. Grassi 74, 20157, Milan, Italy, Cecilia Eugenia Gandolfi, Department of Biomedical Sciences for Health, University of Milan, Via C. Pascal 36, 20133, Milan, Italy. Equally contributing Authors (Elia Biganzoli, Massimo Galli).

**Keywords:** SARS-CoV-2, coronavirus, COVID-19, antibodies, prevalence, rapid test, immunocromatographic test

## Abstract

**Background:** Several municipalities in the Lombardy Region have been affected by the SARS-CoV-2 infection since the earliest stages of the epidemic. To date, 89442 confirmed cases have been diagnosed in Lombardy, and mortality in several municipalities has already surpassed that of the past decade. Currently, the true extent of the SARS-CoV-2 infection remains unknown as several affected subjects may have been asymptomatic or have presented mild disease, thus not resulting in the identified COVID-19 cases.

**Methods:** This cross-sectional study aims to define the spread of infection within the population by determining the seroprevalence of IgG antibodies directed against SARS-CoV-2 by rapid immunochromatographic testing and subsequent confirmation by serology on venous blood by liquid phase immunochemical testing, also allowing to compare the two methods. Testing will be performed on adults and minors residing, domiciled or working in several municipalities of the Lombardy Region, involved in the initial stages of the epidemic. The study will include rapid finger-prick testing and venous sampling for antibodies against SARS-CoV-2, and nasopharyngeal swabbing (NPS). Concurrent notification of test results will occur via the regional healthcare information system (SISS).

**Discussion:** This study was developed with the desire to understand the seroprevalence of SARS-CoV-2 infection and the epidemiological transmission characteristics of this virus. Understanding the spread and severity of the disease could help in the implementation of effective infection surveillance containment and countermeasures facilitating the identification of cases that have been exposed to the virus and the traceability of contacts.

**This study has been approved by the Ethics Committee of the University of Milan (35/2020)**.

## BACKGROUND

Several municipalities of the Lombardy Region have been affected by the Severe Acute Respiratory Syndrome Coronavirus 2 (SARS-CoV-2) infection since the earliest stages of the epidemic. In fact, the first so called “red zone”, a quarantined area where all movements were restricted, was established in the Lodi province (south-eastern Lombardy) on the 23^rd^ of February, following the first Coronavirus Induced Disease (COVID-19) hospitalization from Codogno (LO).

To date, a total of 89442 SARS-CoV-2 cases have been confirmed in the Lombardy Region^1^, however the true extent of the SARS-CoV-2 infection remains unknown. In particular, the number of infected subjects having presented minimal or no symptomatology has not been precisely determined.

The World Health Organization (WHO) recognized the existence of a coronavirus responsible for pneumonia in Wuhan, China on December 29th 2019^2^. This virus, initially identified as n-CoV-2019 (novel Coronavirus 2019), was subsequently referred to as SARS-CoV-2, and its clinical manifestations known as Coronavirus Induced Disease 2019 (COVID-19). The pandemic was declared on March 11th 2020^3^.

Currently, the WHO considers forwarding scientific knowledge on mass screening essential for the battle against the spread of SARS-CoV-2, in order to allow better understanding and planning of current and future containment policies.

All countries that have made extensive use of diagnostic testing on the whole population, regardless of presence or absence of symptoms, have achieved a drastic drop in infection and spread^4^.

To date, the only diagnostic tool to be officially recognized by the Italian Ministry of Health for SARS-CoV-2 infection is the NPS, that uses real-time reverse transcription polymerase chain reaction (rRT-PCR) to identify and amplify traces of the SARS-CoV-2 genome within a patient’s sample^5^. This diagnostic method has several limits, including high costs, long complete analysis times, and many false negatives (low diagnostic sensitivity).

Alongside the diagnostic aspect, the development of knowledge regarding disease spread and prevalence in the population will become increasingly important in the near future. For this reason, the development of research on seroprevalence with appropriate methods is of fundamental importance.

This study was developed with the desire to understand the seroprevalence of SARS-CoV-2 infection and the epidemiological transmission characteristics of this virus in the earliest affected areas in Italy. Understanding the spread and severity of the disease could help in the implementation of effective infection surveillance containment and countermeasures facilitating the identification of cases that have been exposed to the virus and the traceability of contacts.

## METHODS & DESIGN

The primary objective of this cross-sectional study is to determine the spread of infection within the population by determining the seroprevalence of IgG antibodies directed against SARS-CoV-2 by rapid immunochromatographic testing and subsequent confirmation by serology on venous blood by liquid phase immunochemical testing.

Secondary objectives include comparing IgG and IgM antibody positivity obtained by rapid testing via direct viral research to serological data obtained by the immunochemical method. We also aim to establish a database for future prospective projects eg. serial antibody titer assessed over time, possibly expanding the study to several municipalities particularly affected by the SARS-CoV-2 epidemic around the Lombardy region. This may help localize and isolate possible current and future outbreaks.

For the municipalities included in first so-called “red zone”, thus more severely hit, we hypothesize a seroprevalence of IgG antibodies directed against SARS-CoV-2 between 30% and 50%, due to probable unrestricted viral circulation in the days or weeks before the first hospitalized case. For the other, less severely affected, municipalities we estimate a prevalence between 10 and 30%.

According to current regional legislation (DGR n. XI/3114 and DGR n. XI/3131) during the second phase of the epidemic, NPS will be performed in all symptomatic individuals, flagged as “suspect COVID-19 cases” by their family doctor, and in all traced contacts prior to readmission in society. Although rRT-PCR on NPS is the current gold standard diagnostic test for SARS-CoV-2, diagnostic accuracy depends on several pre-analytical and analytical variables, and many patients can be asymptomatic (thus currently will not receive testing in the Lombardy region). The measurement of specific COVID-19 antibodies could serve as an additional tool for disease detection and management, playing a potential key role in surveillance and screening. The latter may be even more advantageous if implemented in the form of rapid finger-prick testing. In fact, as mentioned above, one of our secondary objectives is the comparison of IgG and IgM antibody positivity obtained by rapid testing, to serological data obtained by the immunochemical method. We hypothesize that antibody detection by rapid finger-prick testing will be non-inferior to detection by venous sampling.

Testing will be performed at specific locations in the various municipalities, equipped with the following rooms:

- A waiting room of suitable size to allow the simultaneous presence of 8-10 evenly spaced (at least 1.8 m apart) individuals equipped with suitable PPE (surgical masks).
- A room for collection of personal data and for carrying out rapid tests.
- A room for the execution of venous blood samples.
- A room for carrying out NPS.
- A room for the dressing and undressing of healthcare workers.
- A storage room with refrigerator.

### Inclusion and exclusion criteria

All following subjects may be included in the study:

1. The entire adult population living in the municipality, including residential care facilities.
2. All individuals over 18 who were present in the municipality on a daily basis for work reasons during the epidemic eg. law enforcement officers, traders, workers in local companies etc.
3. All minors residing and/or domiciled in the municipality, upon explicit parental/guardian request.

An absolute exclusion criterion is represented by being quarantined. However, data regarding quarantine will be taken into account for defining the dynamics of the epidemic.

### Population follow-up and counseling

1. Individuals with **negative** results to rapid testing who do not present symptoms suggestive for acute infection will be sent home, after counseling on appropriate behaviors aimed at avoiding infection.
2. Individuals with **negative** results to rapid testing who, however, present symptoms suggestive for acute infection will be evaluated by medical personnel to determine the need for further investigations (imaging or ER access) and quarantined, at home or in a different location if home is deemed unsuitable, pending swab results. These individuals will receive extensive counseling on best-practices during the quarantine period.
3. Individuals with **positive** results to IgM rapid testing, regardless of IgG results, will be quarantined at home, or in a different location if home is deemed unsuitable, pending swab results. If swab results are also positive, quarantine will be confirmed and extended and counseling on best-practices during the quarantine period will be provided.
4. Individuals with **positive** results to IgG rapid testing only will be sent home, without need for fiduciary quarantine.
5. Quarantined individuals will be provided with pulse oximeter, surgical masks and extensive counseling on best-practices during the quarantine period.
6. All **swab positive** individuals, regardless of rapid test results, will be notified to local health authorities using a specific infectious disease digital notification platform for the Lombardy Region. The swab positive individual will thus be quarantined, and follow-up will be carried out by the local health authorities, as indicated in current regional protocols.

### Results reporting

Rapid test results will be communicated in real time and a full report comprising collected anamnestic and epidemiological data, and rapid test results will be emailed or printed, according to patient preference.

NPS results will be communicated as follows:

- In case of a negative result, attending medical personnel will call the patient directly.
- In case of a positive result, attending medical personnel will call the patient’s family doctor, who will then notify the local health authorities via the regional infectious disease digital notification platform, and communicate the positive result to the patient.

### Sample management

1. Rapid tests will be analyzed by assigned staff within the timeframe suggested by the manufacturer and will then be suitably disposed of by the same staff (biohazardous waste).
2. Venous blood samples will be stored on site, refrigerated at +4°C, and transported to the lab for analysis, daily.
3. Swabs will be stored on site, refrigerated at +4°C, and transported to the lab for analysis, daily (or within 48 hours during the weekend). NPSs are placed in a triple container during transportation: the collection tube is placed in a conical tube (“falcon” type tube), and then in a spill proof box.

### Sampling plan and determination of sample size

For the purposes of the primary objective of the study, random sampling will be carried out from the municipal registry list, stratifying by gender and conventional age classes (0-19; 20-39; 40-59; >=60). At present, geographical stratification does not seem necessary given the homogenous distribution of cases identified to date. Confirmed cases that have already been identified in the municipality must be included in the study sample with the same probability as other subjects. In fact, the rapid test and venous sample seroprevalence results on previously confirmed cases will provide further data useful for comparison.

A sample size of 355 subjects from a population of 4616 individuals provides a bilateral 95% confidence interval with an accuracy (half-amplitude) of 0.05 when actual prevalence is close to 0.5^6^. In case of higher or lower prevalence, the sample size decreases to 302 subjects in case of prevalence equaling 0.3, very close to current estimates, and to 234 subjects in case of a more skeptical prevalence hypothesis of 0.8. Considering the largest population sample combined to a possible 20% drop-out, sample size increases to 444 subjects.

Assuming an infinite population^7,8^, sample size would be determined at 402 subjects, to be increased to 503 subjects considering 20% drop-out. Thus, a sample size of around 500 subjects would seem reasonable for the main study objective.

Regarding the secondary objective, relating to sensitivity and specificity of the rapid test compared to the gold-standard on venous blood samples, the sample size required for a bilateral 95% confidence interval with a maximum amplitude of 0.1, is 527 subjects for sensitivity and 438 subjects for specificity, assuming a prevalence of 0.3 with optimal sensitivity and specificity equal to 0.9. Thus, in order for both confidence intervals to have a maximum amplitude of 0.1, the largest of the two sample sizes (527 subjects) must be chosen^9^. Also, sample size should be further increased to 659 subjects to allow for 20% drop-out. Accordingly, in case of hypothesized sensitivity and specificity values equal to 0.8, the sample size should be increased to 880 subjects, and to 1100 subjects to include 20% drop-out. However, in an optimal situation, the sample size required for the secondary objective appears compatible with that needed for the primary objective. In fact, a study that could effectively recruit at least 500 people would continue to offer expected amplitudes of confidence intervals up to 0.144 for sensitivity and <0.1 for specificity, even in conditions of lower diagnostic performance, always hypothesizing a prevalence of 0.3.

Sample size calculations were carried out using PASS v. 16.0.3 validated software.

## DISCUSSION

### Location and testing procedures

Testing will be performed at various locations in the selected municipalities, equipped with the following rooms:

- A waiting room of suitable size to allow the simultaneous presence of 8-10 evenly
- spaced (at least 1.8 m apart) individuals equipped with suitable PPE (surgical masks).
- A room for collection of personal data and performing rapid tests.
- A room for the execution of venous blood samples.
- A room for carrying out NPS.
- A room for the dressing and undressing of healthcare workers.
- A storage room with refrigerator.

Separate paths will be defined for the entrance and exit of tested subjects, particularly to prevent loitering between individuals waiting to be tested and those already tested, as well as to prevent large gatherings in confined spaces like corridors, making social distancing impracticable.

Subjects are tested as follows:

1. Assembly of up to 10 people every 30 minutes to be examined according to geographical location. Participation in the study is voluntary. Those called upon must be equipped with a correctly worn surgical mask. Security personnel will be assigned to ensure correct use of PPE and compliance with safety distance.
2. Acquisition of informed consent.
3. Rapid testing for antibodies against SARS-CoV-2 in all individuals in the assembled group.
4. Anamnestic and personal data collection on a local network electronic database and printing of labels for sample identification.
5. Venous blood sampling in all those resulting positive to the rapid test and, at random, in a proportion of negative subjects, still to be determined.
6. Swabbing of all individuals resulting positive to the rapid test, all people with recent history of symptoms compatible with COVID-19, and in a proportion of negative subjects, yet to be determined, chosen at random.

Given that examining the largest number of people in the shortest time possible is of paramount importance in a cross-sectional study, we decided to perform rapid testing as the first procedure, and to gather epidemiological and anamnestic data while waiting for the appropriate reading time (10 to 20 minutes). This will enable us to perform between 200 and 250 rapid tests per day.

### Personnel protection measures

Seeing as the SARS-CoV-2 epidemic is still active in the Lombardy region, participant and personnel safety will be a main concern. Personal protective equipment (PPE) and sanitization of all rooms and equipment will be guaranteed for the all duration of the study.

All patients must be equipped with a correctly worn surgical mask and disposable gloves. PPE will be provided according to personnel role, as follows:

1. Surgical mask and disposable gown for personnel assigned to anamnestic, personal data, and rapid test collection
2. FFP2 or equivalent filtered mask, surgical cap, visor, gloves and waterproof disposable gown for personnel assigned to NPS collection and venous sample collection.

Moreover, we decided to position a plexiglass barrier with a specifically-designed hole through which the rapid test is performed. This guarantees additional safety for personnel assigned to rapid testing and data collection, without the discomfort of wearing PPEs. The same barrier was proposed to the nurses assigned to venous blood sampling, but was not implemented based on previous negative experiences with this method of blood collection through the plexiglass barrier.

Rooms and surfaces will be sanitized three times a day using 0.1-0.5% sodium hypochlorite solution for large surfaces eg. floors, walls, chairs; and 62-71% alcohol for small surfaces (whenever there is biological contamination) and remaining instrumentation.

Sanitization will be carried out with disposable cloths or paper towels, to be suitably eliminated (biohazardous waste).

Sanitization of any medical devices (sphygmomanometer, pulse oximeter, etc.) will be carried out between each use.

### Data management

Study data will initially be stored on a certified database of the i-Care platform, provided by MEDISPA. Subsequently, an original database will be created, in collaboration with the University of Milan Data Science Research Center, for data processing and analysis, according to i-Care database data protection requirements, from which data transfer will take place. Patient data will be traceable to a completely anonymous identification code (no data from which patient identity could be derived will be recorded), thus only the doctor assigned to patient data collection and, if required, subjects authorized by national privacy regulations will be able to trace data back to the specific patient via the abovementioned identification code. In any case, these subjects are bound by confidentiality according to current legislation. Patient data will be made public only via scientific publications or conferences, and only in anonymous aggregate form, always respecting current privacy laws (including but not limited to, “*legge privacy*”, 18/05/2018, n. 51 Legislative Decree; and the EU (GDPR) 2016/679^10^.

The protection of individuals will also be guaranteed according to the Oviedo Convention^11^ and the Declaration of Helsinki regarding ethical principles for medical research involving human subjects^12^, adopted by the World Medical Association in 1964.

## Data Availability

The article is reporting the protocol for the study. Data will be available as soon as the study will be completed

## LIST OF ABBREVIATIONS

SARS-CoV-2: severe acute respiratory syndrome coronavirus 2
COVID-19: COronaVirus Induced Disease 2019
rRT-PCR: real-time reverse transcription polymerase chain reaction
NPS: nasopharyngeal swab
SISS: Sistema Informativo SocioSanitario (regional healthcare information system)
WHO: World Health Organization
PPE: personal protective equipment

## DECLARATIONS

### Ethics approval and consent to participate

The study have been performed in accordance with the Declaration of Helsinki and was approved by the Ethics Committee of the University of Milan on the 21^st^ of April 2020 (reference number 35/2020). Informed consent to participate in the study will be obtained in written form from all participants, or their parent or legal guardian in case of minors.

### Consent for publication

Not applicable.

### Availability of data and materials

The datasets used and analysed during the current study are available from the corresponding author on reasonable request.

### Funding

This study will be carried out with the non-conditioning contribution of CISOM, Fondazione

Rava, Armani C/O Unimi e Mediolanum C/O Fbf/Sacco.

Vittore Scolari received support from the LabEx DEEP (ANR-11-LABX-0044, ANR-10-427 IDEX-0001-02)

### Authors’ contributions

MG, GP, DB, AG and FC contributed to the conception and design of the protocol and revised the manuscript;

EB and PB contributed to the design of the protocol, the acquisition, analysis and interpretation of data, and revision of the work;

RR and VS contributed to the acquisition, analysis and interpretation of data;

CEG and SC drafted and substantially revised the work.

All authors declare to approve the submitted version and agree to be personally accountable for their contributions.

## Acknowledgements

The authors would like to acknowledge the substantial contribution provided by Medispa s.r.l., which contributed to the protocol by providing expertise, personnel and the informatic platform.

